# Modelling the situation of COVID-19 and effects of different containment strategies in China with dynamic differential equations and parameters estimation

**DOI:** 10.1101/2020.03.09.20033498

**Authors:** Xiuli Liu, Geoffrey Hewings, Shouyang Wang, Minghui Qin, Xin Xiang, Shan Zheng, Xuefeng Li

**Affiliations:** Academy of Mathematics and Systems Science, Chinese Academy of Sciences, Zhongguancun East Road No. 55, Beijing 100190, People’s Republic of China; University of Chinese Academy of Sciences, Beijing, 19 A Yuquan Road, Shijingshan District, Beijing 100049, People’s Republic of China; Center for Forecasting Science, Chinese Academy of Sciences, Zhongguancun East Road No. 55, Beijing 100190, People’s Republic of China; Regional Economics Applications Laboratory, University of Illinois, 1301 W Gregory #236, Urbana, IL 61801, USA

**Keywords:** dynamically modeling, parameters estimation, sensitive analysis, effects of different containment strategies, novel coronavirus (COVID-19)

## Abstract

This paper proposed a quarantine-susceptible-exposed-infectious-resistant (QSEIR) model which considers the unprecedented strict quarantine measures in almost the whole of China to resist the epidemic. We estimated model parameters from published information with the statistical method and stochastic simulation, we found the parameters that achieved the best simulation test result. The next stage involved quantitative predictions of future epidemic developments based on different containment strategies with the QSEIR model, focused on the sensitivity of the outcomes to different parameter choices in mainland China. The main results are as follows. If the strict quarantine measures are being retained, the peak value of confirmed cases would be in the range of [52438, 64090] and the peak date would be expected in the range February 7 to February 19, 2020. During March18-30, 2020, the epidemic would be controlled. The end date would be in the period from August 20 to September 1, 2020. With 80% probability, our prediction on the peak date was 4 days ahead of the real date, the prediction error of the peak value is 0.43%, both estimates are much closer to the observed values compared with published studies. The sensitive analysis indicated that the quarantine measures (or with vaccination) are the most effective containment strategy to control the epidemic, followed by measures to increase the cured rate (like finding special medicine). The long-term simulation result and sensitive analysis in mainland China showed that the QSEIR model is stable and can be empirically validated. It is suggested that the QSEIR model can be applied to predict the development trend of the epidemic in other regions or countries in the world. In mainland China, the quarantine measures can’t be relaxed before the end of March 2020. China can fully resume production with appropriate anti-epidemic measures beginning in early April 2020. The results of this study also implied that other countries now facing the epidemic outbreaks should act more decisively and take in time quarantine measures though it may have negative short-term public and economic consequences.

## Introduction

In late December, 2019, an atypical pneumonia case, caused by a virus called COVID-19, was first reported and confirmed in Wuhan, China. Although the initial cases were considered to be associated with the Huanan Seafood Market, the source of the COVID-19 is still unknown. The confirmed cases increased with exponential speed, from 41 on January 10, 2020 to 5,974 on January 28, 2020 in mainland China, far exceeding those of the SARS epidemic in 2003 (see figure 1). By February 22(24:00 GMT), 2020, there have been 76,936 cumulative confirmed cases of COVID-19 infections in mainland China, including 2,442 cumulative deaths and 22,888 cumulative cured cases. 64,084 cumulative confirmed cases were in Wuhan, accounting for 83.3% of the cumulative confirmed cases in mainland China. Equally of concern, a WHO news release noted that 1,400 cases were reported in 26 countries outside China, with the Republic of Korea (346), Japan (105) and Singapore (86) ranked as the top 3 (figure 2), while 35 cases were reported in United States of America1.

**Figure 1.**
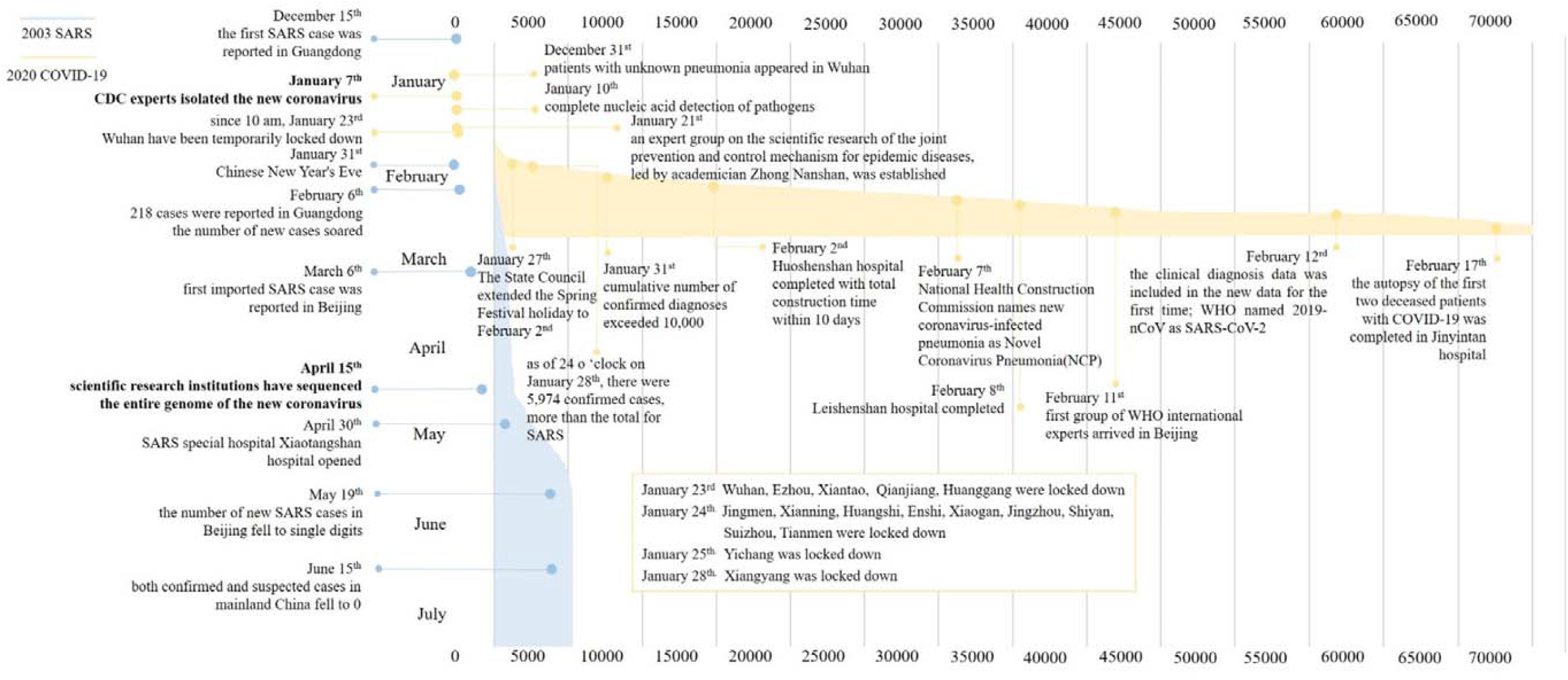
Timeline comparison between SARS and COVID-19

**Figure 2.**
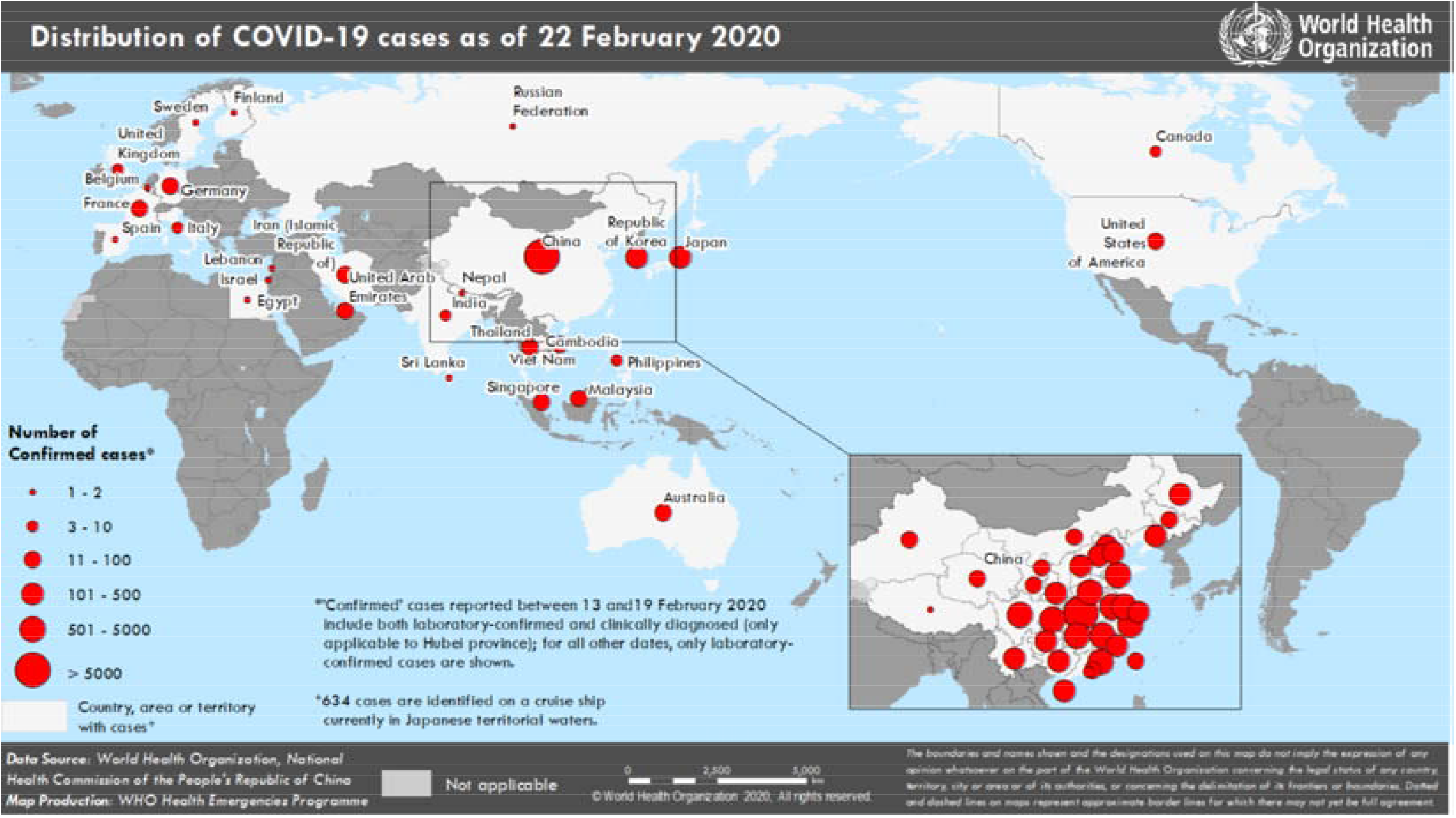
Countries, territories or areas with reported confirmed cases of COVID-19, 22 February 2020 (sourced from WHO) *The situation report includes information provided by national authorities as of 10 AM Central European Time

The transmissibility of COVID-19 — or at least its geographical distribution (figure 2) — seems to be higher and broader than initially expected (Horton, 2020). Compared to SARS-CoV (9.56% mortality) and MERS-CoV (34.4% mortality), the COVID-19 appears to be less virulent at this point except for the elderly and those with underlying health conditions (table 1). COVID-19 was confirmed as subject to human-to-human transmission and it is very contagious. The basic reproduction number R0 for COVID-19 was estimated by WHO and some research institutes in the range of 1.4-6.6 (table 2). This value is slightly higher than that of the 2003 SARS epidemic, and much higher than that of influenza and Ebola. The incubation days of COVID-19 in Wuhan city is 5-10 days with a mean of 7 days (Fan *et al*., 2020). On average, the duration from confirmed stage to cure or death is 10 days in nation-wide reporting according to Guan *et al*. (2020). A long incubation period and an associated large number of patients with mild symptoms increase the difficulty of prevention and control of the epidemic. The likelihood of travel-related risks of the disease spreading has been noted by Bogoch *et al*. (2020) and Cao *et al*. (2020a) wherein they indicated the potentials for further regional and global spread (Leung *et al*., 2020).

**Table 1.**
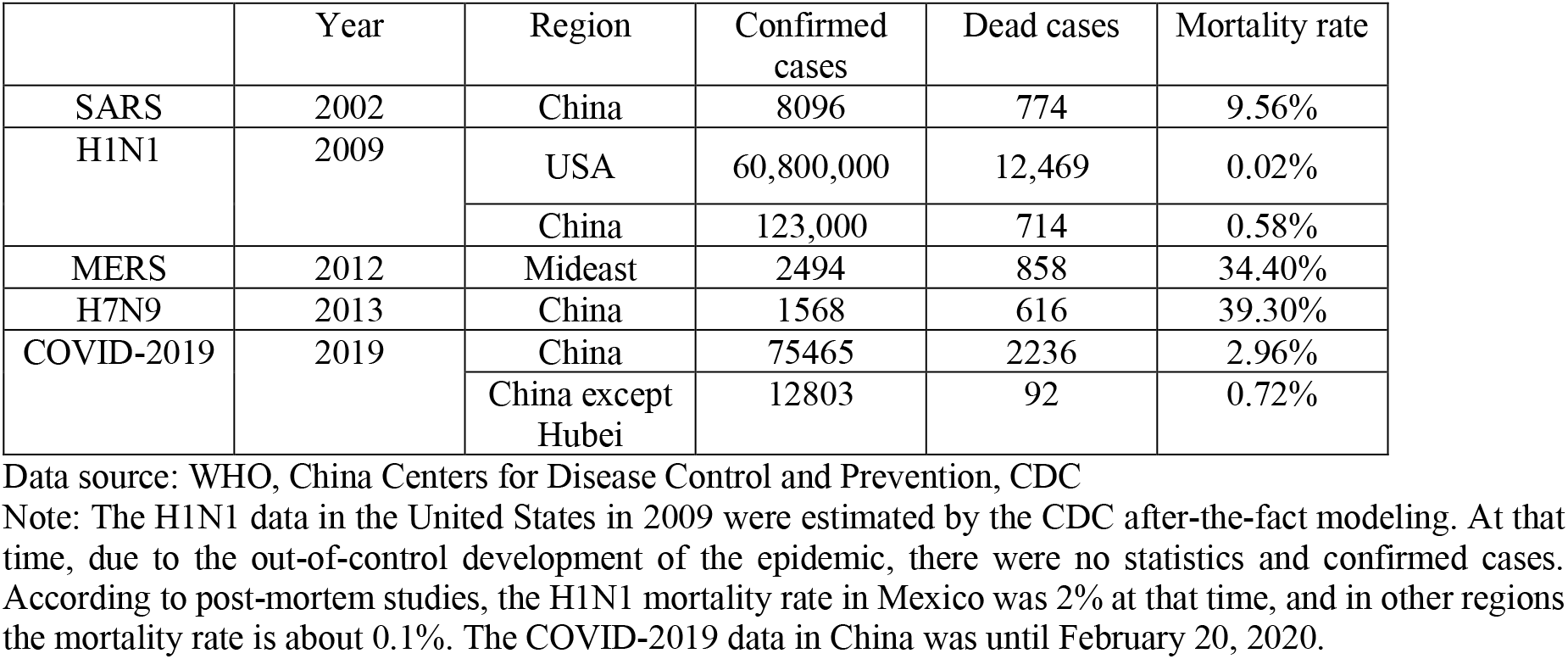
Several major outbreaks in the world over the past 20 years

**Table 2.** The summary table of the estimated basic reproduction number R0 of four epidemics from different studies

| Epidemic | R0 | Sourced studies |
| --- | --- | --- |
| COVID-19 | 1.4-2.5 | WHO(1.4-2.5); Hermanowicz (2020)(2.4-2.5); Cowling and Leung (2020)(2.2); Riou and Althaus (2020)(2.2); Li *et al.* (2020a)(2.2); Rabajante (2020)(2.0); Su *et al.* (2020)(2.24-3.58); Li *et al.*, (2020c)( 2.2-3.1); Geng *et al.* (2020)(2.38-2.72) |
|  | 2.5-3.0 | Zhou *et al.* (2020) (2.8-3.9); Su *et al.* (2020) (2.24-3.58); Geng *et al.* (2020) (2.38-2.72); Xiong and Yan (2020) (2.7); Li *et al.*, (2020c) (2.2-3.1); Wu *et al.* (2020a) (2.68) |
|  | 3.0-3.5 | Zhou *et al.* (2020) (2.8-3.9); Su *et al.* (2020) (2.24-3.58); Li *et al.*, (2020c) (2.2-3.1); Liu *et al.* (2020c) (3.28); Cao *et al.* (2020b) (3.24); Read *et al.* (2020) (3.11); Cao *et al.* (2020a) (3.24) |
|  | 3.5-4.0 | Zhou *et al.* (2020) (2.8-3.9); Su *et al.* (2020) (2.24-3.58); Zhang *et al.* (2020) (3.6) |
|  | 4.0-7.0 | Shen *et al.* (2002) (4.71); Sanche *et al.* (2020) (4.7-6.6) |
| SARS | 2.0-5.0 | Wallinga and Teunis (2004) |
| Ebola | 1.5-2.5 | Althaus (2014) |
| influenza | 2.0-3.0 | Mills *et al.* (2004) |

As the epidemic broke out on the eve of the Spring Festival, large-scale population movements and gatherings of people aggravated the epidemic. After the outbreak, local governments have adopted a series of unprecedented mitigation policies in place to contain the spread of the epidemic. The major local public emergency started with a category Class I response to health incidents, with positively diagnosed cases either quarantine or put under a form of self-quarantine at home (Gan *et al*., 2020). Suspicious cases were confined in monitored house arrest. Most exits and entries into cities were shut down. Certain categories of contact were banned; for instance, universities and schools remained closed, and many businesses remained closed. People were asked to remain in their homes for as much time as possible (Fahrion *et al*., 2020). These interventions have reduced the population’s contacts to a certain extent, helped to cut off pathways for the spread of the virus and reduce the rate of disease transmission.

However, the long-term management and control has brought considerable inconvenience to the daily lives of people. The failure of factories to start on time and run normally after the Spring Festival also had severe effects on Chinese national and global economies. Ayittey *et al*. (2020) and CNN Business (2020) estimated it would result in China’s GDP declining 4.5% year-on-year in Q1 in 2020; the loss in China would be up to $62 billion in the same quarter. Zhang (2020), Huang (2020), Li and Zhang (2020) and IMF News (2020) considered the growth of China’s GDP would be 5.0%-5.6% in 2020, decrease 0.5-1.1 percentage points from 2019. IHS Markit (2020) estimated a reduction of global real GDP of 0.8% in Q1 and 0.5% in Q2 in 2020, and the global real GDP would be reduced by 0.4% in 2020. The longer the duration of the epidemic, the more negative the impacts on China and the rest of the world, with the latter effects largely centered on disruptions in increasingly complicated supply chains. Therefore, it is important to estimate the dynamic evolution mechanism of the epidemic in mainland China, to find when the epidemic will end and how this result depends on different containment strategies. These are issues of great significance with important clinical and policy implications (Joseph *et al*., 2020).

## QSEIR Model

The traditional infectious disease dynamics susceptible–exposed–infectious–resistant (SEIR) model has been very popular in analyzing and predicting the development of an epidemic (see Lipsitch *et al*., 2003; Pastor-Satorras, 2015). SEIR models the flows of people between four states: susceptible (*S*), exposed (*E*), infected (*I*), and resistant (*R*). Each of those variables represents the number of people in those groups. Assume that the average number of exposed cases that are generated by one infected person of COVID-19 is *β*. The parameter *β* is similar to the basic reproduction number which can be thought of as the expected number of cases directly generated by one case in a population where all individuals are susceptible to infection. Considering the protective measures were taken, *β* should be smaller than the basic reproduction number in table 2. An individual in the exposed state (type E) will have the probability *δ* changes to individuals in the infected state (type I), and an individual in the infected state (type I) will change to the cure state (type R) with a probability of *γ* or to death state (type F) with a probability of *η* per unit time. In contrast to the traditional SEIR model, we propose a quarantine-susceptible-exposed-infectious-resistant (QSEIR) model that considers the unprecedented strict quarantine measures in mainland China to resist the epidemic. The parameter, *α*(*t*), was designed to represent the ratio of people who was not restricted to a specific area and had chances to contact with COVID-19 virus during special period. The *α*(*t*) and *β*(*t*) vary according to the strength of the prevention and control measures for the epidemic. To make the model accord with reality, contrast with the standard SEIR model, we added two parameters *Δ*(*t*) and *θ*(*t*). The *Δ*(*t*) is the ratio of people with vaccination at time t. *θ*(*t*) is the natural mortality of the population in a region at time t (figure 3). The value of *δ*(*t*) is closely related with the virus incubation and infectious periods and *γ*(*t*) is dependent on the treatment level and patients’ health status. It is assumed that the virus incubation period is 7 days and the duration from confirmed stage to cure or death is 10 days based on nation-wide information (Guan *et al*., 2020; Fan *et al*., 2020). The model is an ordinary differential equation model, described by the following equation.

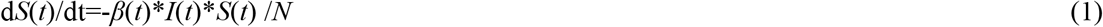

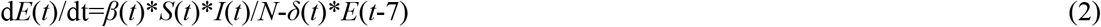

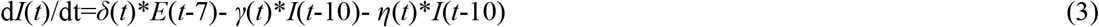

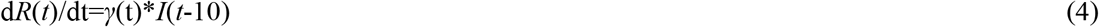

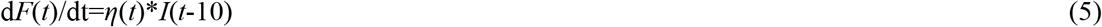

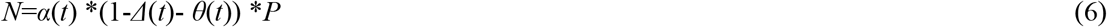

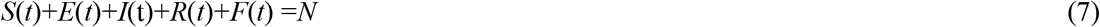

**Figure 3.**
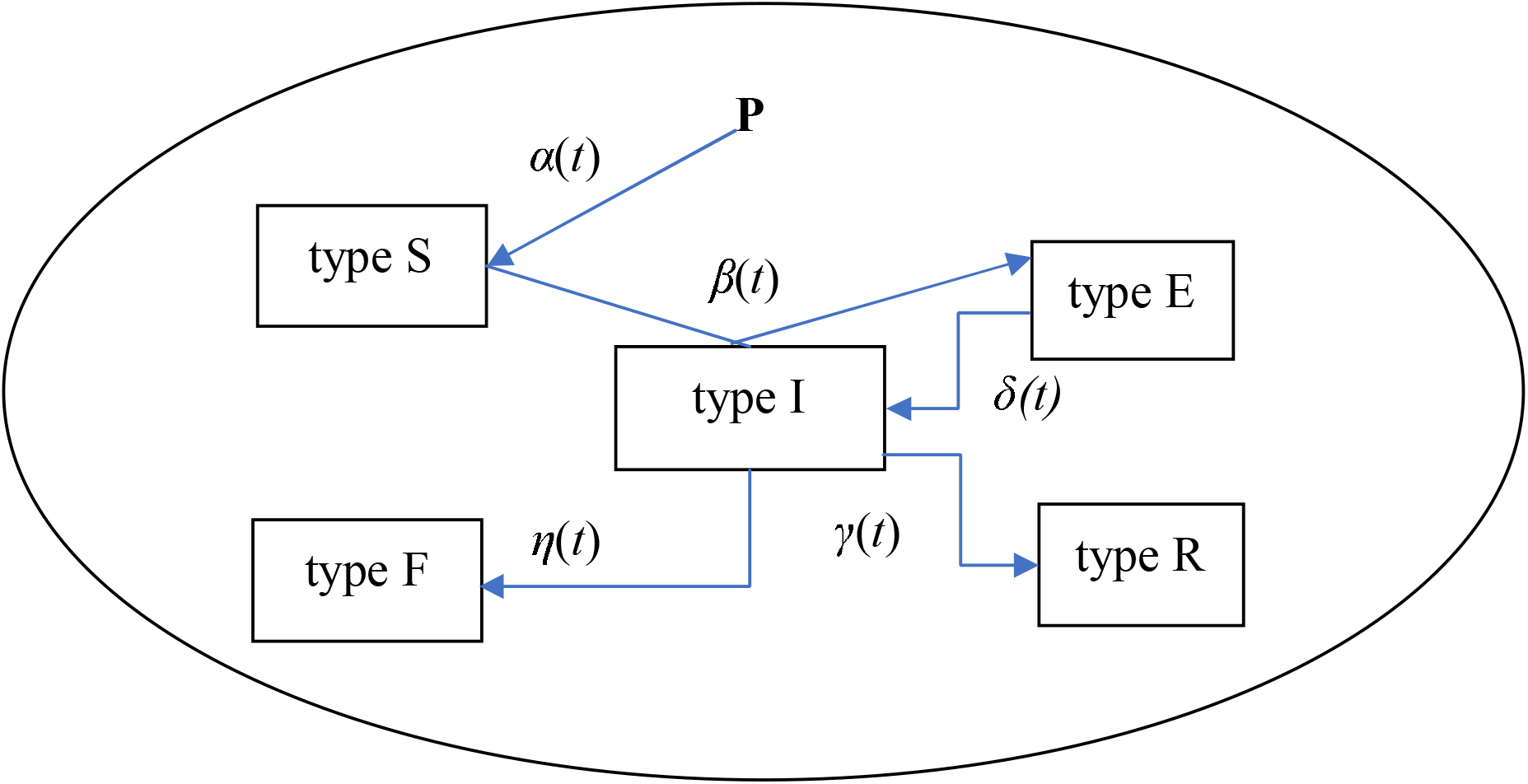
The changes of people among different status when one epidemic outbreaks

Equation (6) is specially designed to fit for China’s actual epidemic prevention measures. In actual calculations, *Δ*(*t*) was assumed to be 0, because no vaccination has yet been developed. *θ*(*t*) can also be assumed to be 0 if we are only concerned with the fatality of CONVID-19. The other four parameters *β*(*t*), *γ*(*t*), *δ*(*t*) and *η*(*t*) are not easy to determine, since the virus incubation period, infectious period, and case statistics that have close relationships with these parameters have varying (unknown) degrees of accuracy. The choice of estimation techniques for the key epidemiological parameters in the QSEIR model of COVID-19 has become a research priority (Cao *et al*., 2020b).

### Data Source

We obtained the number of COVID-19 cases time series data from January 10 to February 22, 2020 for mainland China released by the National Health Commission of China and health commissions at the provincial level in China2. Due to limited testing and treatment resources while facing a major outbreak with a sudden onset, there was under-screening and under-reporting in the early stages of the epidemic in its epicenter, Wuhan, and this generated biases in the data during the early stages (Cao *et al*., 2020b). Note that this challenge also existed in SARS and other coronavirus outbreaks (Hartley and Smith, 2003; Razum and Becher, 2003). After the isolation of Wuhan on January 23, 2020 with the stricter requirements of data statistics and the provision of detection levels, the data are more and more reliable.

### Parameters Estimation

We estimated model parameters reversely with QSEIR model by equations (8)-(12). *β*(*t*), *γ*(*t*), *η*(*t*) and *δ*(*t*) can be calculated (see table 4).

**Table 3.** The predicted peak date of the confirmed cases in 2020 in mainland China from different studies

|  |  |  |  |  |  |
| --- | --- | --- | --- | --- | --- |
| Jan. 14 | Feb.7 | Feb.9 | Feb.11 | Feb.13 | Feb.12-13 |
| Yan *et al.* (2020) | Tang *et al.* (2020) | Batista (2020) | Tomie (2020) | Wang *et al.* (2020b) | Gamero *et al.* (2020) |
| Feb.16 | Feb.16/17 | Mid of Feb. | Feb.19 | Feb.29 | The beginning of March |
| Xiong and Yan (2020) | Li *et al.* (2020a) | Hermanowicz (2020) | Shi *et al.* (2020) | Anastassopoulou *et al.* (2020) | Geng *et al.* (2020) |

**Table 4.** The calculated value of four parameters with equations (8)-(12)

| Date in 2020 | $\beta(t)$ | $\gamma(t)$ | $\delta(t)$ | $\eta(t)$ |
| --- | --- | --- | --- | --- |
| Jan.23 | 0.0616 | 0.1212 | 0.6000 | 0.2424 |
| Jan.24 | 0.0954 | 0.1481 | 19.0417 | 0.5926 |
| Jan.25 | 0.1458 | 0.3929 | 25.4815 | 0.5357 |
| Jan.26 | 0.1965 | 0.0488 | 14.2407 | 0.5854 |
| Jan.27 | 0.3179 | 0.0957 | 13.0221 | 0.2766 |
| Jan.28 | 0.4043 | 0.2529 | 3.7125 | 0.1529 |
| Jan.29 | 0.4978 | 0.0894 | 1.6203 | 0.1617 |
| Jan.30 | 0.5911 | 0.1090 | 1.0081 | 0.0998 |
| Jan.31 | 0.6778 | 0.1300 | 0.7820 | 0.0830 |
| Feb.1 | 0.7847 | 0.1102 | 0.4468 | 0.0584 |
| Feb.2 | 0.8778 | 0.1217 | 0.4051 | 0.0472 |
| Feb.3 | 0.9716 | 0.0840 | 0.3499 | 0.0342 |
| Feb.4 | 1.0857 | 0.0995 | 0.3194 | 0.0249 |
| Feb.5 | 1.1482 | 0.0600 | 0.2424 | 0.0168 |
| Feb.6 | 1.1659 | 0.0674 | 0.1747 | 0.0127 |
| Feb.7 | 1.1719 | 0.0688 | 0.1732 | 0.0116 |
| Feb.8 | 1.1494 | 0.0644 | 0.1230 | 0.0096 |
| Feb.9 | 1.2870 | 0.0560 | 0.1281 | 0.0086 |
| Feb.10 | 1.3310 | 0.0520 | 0.1061 | 0.0079 |
| Feb.11 | 1.4721 | 0.0455 | 0.0816 | 0.0059 |
| Feb.12 | 1.5232 | 0.0604 | 0.5748 | 0.0131 |
| Feb.13 | 1.5879 | 0.0354 | 0.1463 | 0.0006 |
| Feb.14 | 1.5590 | 0.0522 | 0.0913 | 0.0054 |
| Feb.15 | 1.5219 | 0.0456 | 0.0851 | 0.0049 |
| Feb.16 | 1.4908 | 0.0448 | 0.0945 | 0.0033 |
| Feb.17 | 1.4570 | 0.0506 | 0.1175 | 0.0029 |
| Feb.18 | 1.4205 | 0.0507 | 0.1302 | 0.0038 |
| Feb.19 | 1.3639 | 0.0473 | 0.0389 | 0.0030 |
| Feb.20 | 1.2854 | 0.0544 | 0.0991 | 0.0030 |
| Feb.21 | 1.2249 | 0.0456 | 0.0998 | 0.0021 |
| Feb.22 | 1.2073 | 0.0400 | 0.0892 | 0.0017 |

From equations (1)-(6), we obtain:

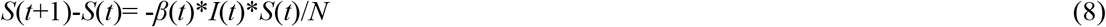

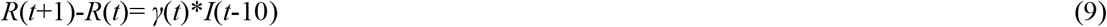

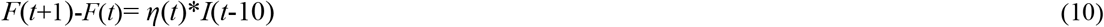

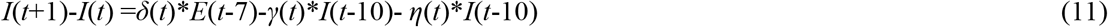

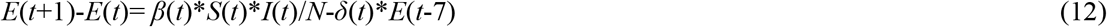

Note that we found some *δ*(*t*) in table 4 was>1, which is obviously incorrect, the reason was mainly because biases in the data during the early stages (Cao *et al*., 2020b). We deleted these data and calculated the average, median and variance of the rest value of the four parameters in first step. In step 2, we deleted values>1.5 times of the column average. In step 3, we calculated the average, median and variance of the rest value of the four parameters (see table 5). With table 5, we set the four parameters belong to the range of their average/median±variance. The parameter *α*(*t*) was roughly estimated as 1.2-2.0 times of cumulative confirmed cases on February 22, 2020 divided by population in mainland China.

**Table 5.** After 3 steps of selection of parameters in Table 4, the statistical characteristic and the estimated values of them

| | $\beta(t)$ | $\gamma(t)$ | $\delta(t)$ | $\eta(t)$ | $\alpha(t)$ |
| --- | --- | --- | --- | --- | --- |
| average | 1.1629 | 0.0562 | 0.2100 | 0.0075 | 0.000076<br>93 |
| median | 0.8778 | 0.0521 | 0.1732 | 0.0054 | 0.000065<br>94 |
| variance | 0.0567 | 0.0002 | 0.0107 | 0.00004 | 8.4553E-10 |
| estimated<br>values | 1.1601 | 0.0509 | 0.2101 | 0.0050 | 0.000069<br>75 |

Then, we set the values of these parameters in their ranges randomly, and input them to QSEIR model, we got *E*(*i*), *I*(*i*), *R*(*i*), *F*(*i*) at each day *i*, we used the real data *I0, E0, R0* and *F0* from February 13 to February 22 in 2020 to test the accuracy of the simulation by *errc*_*k*_ with equation (13).

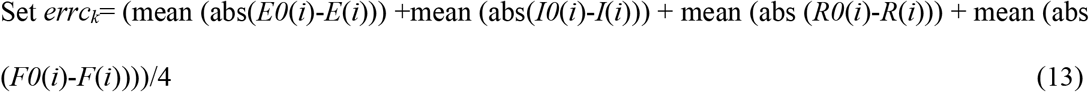

In equation (13), we first calculated the average of the absolute differences between the real data of *E0, I0, R0* and *F0* and their simulated value of *E, I, R* and *F* of *k*th simulation, then we added the four-average value.

5,0000 times simulation were made (figure 4), the result is convergence. The minimum value of *errc*_*k*_ is 20.42% (figure 4), and the estimated values of the five parameters in this case were listed in the last row of table 5. They were applied in the long-term simulation. The minimum value of *errc*_*k*_ is 20.42%, with the current published data that was available, we can use these parameters that can make QSEIR model results with about 80% simulation accuracy.

**Figure 4.**
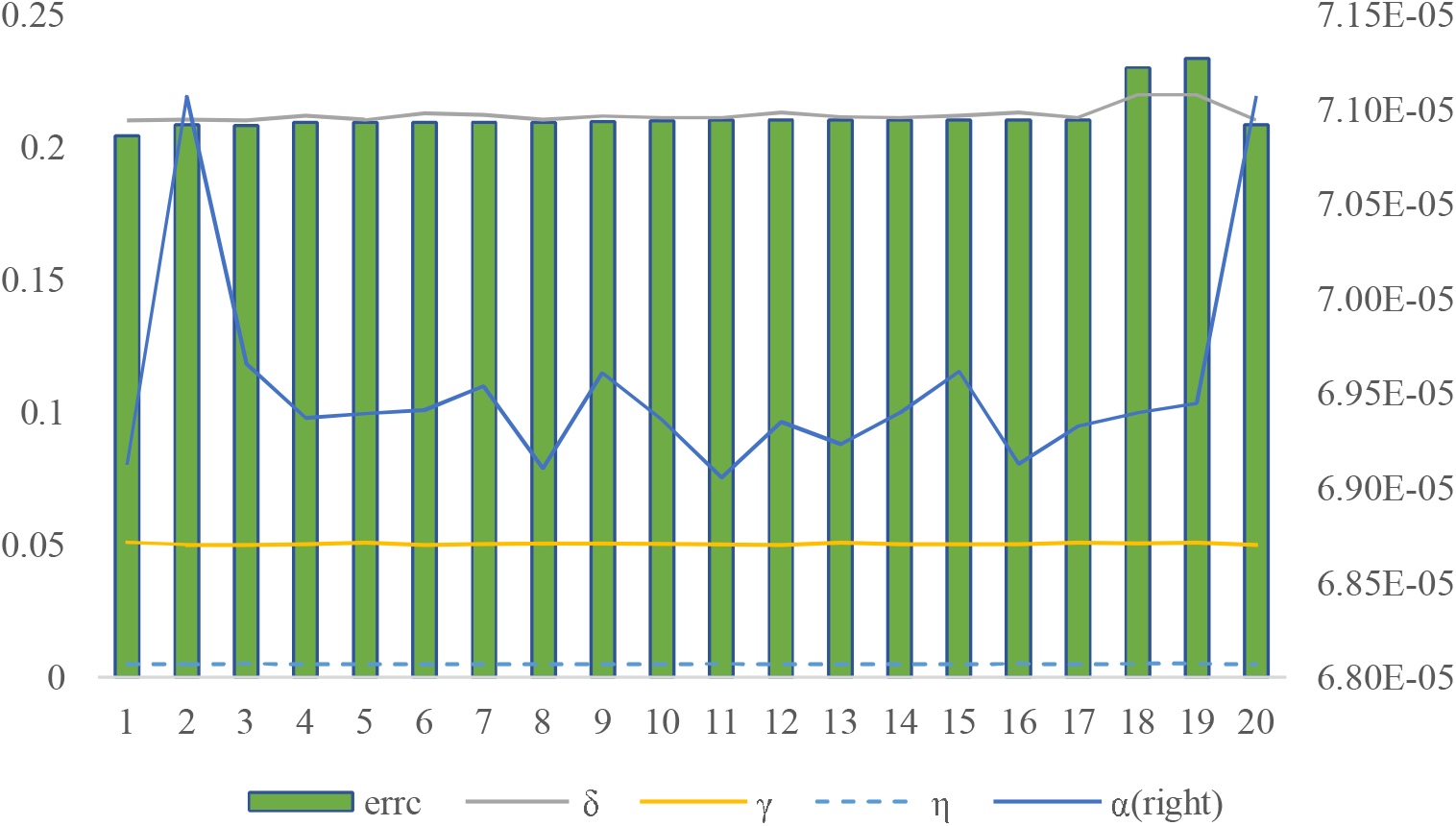
Part of simulation results when parameters were set with random values in their statistical ranges

## Results

We set January 23, 2020 as the beginning date of the simulation; the initial values of variables were set as of this date (table 6). If we set the simulation period *D* as 300 days, input the best parameters we found, with the MATLAB program of QSEIR model, we can present the results shown in figure 5. The results showed that with 80% probability, the peak value of *I* was 58,264 on February 13, 2020. After June 19, 2020, the value of *I* would be < 50 and from July 29, 2020, the number would be smaller than 5. By August 26, 2020, *I* would be smaller than 1, implying that the COVID-19 would essentially end. From March 17, 2020, *E* would be < 5 and, a week later on March 24, the number of *E* would be < 1, which means the epidemic would be totally controlled since this day, no new infected people would appear. The cumulative confirmed cases of COVID-19 in mainland China was estimated to be 97,653, and the cumulative number of deaths was estimated to be 8,754.

**Table 6.** Variables and their initial values in the baseline of QSEIR model

| Variables | $P$ | $E_0$ | $I_0$ | $R_0$ | $F_0$ |
| --- | --- | --- | --- | --- | --- |
| initial values | 14,0005,0000 | 1072 | 771 | 34 | 25 |

**Figure 5.**
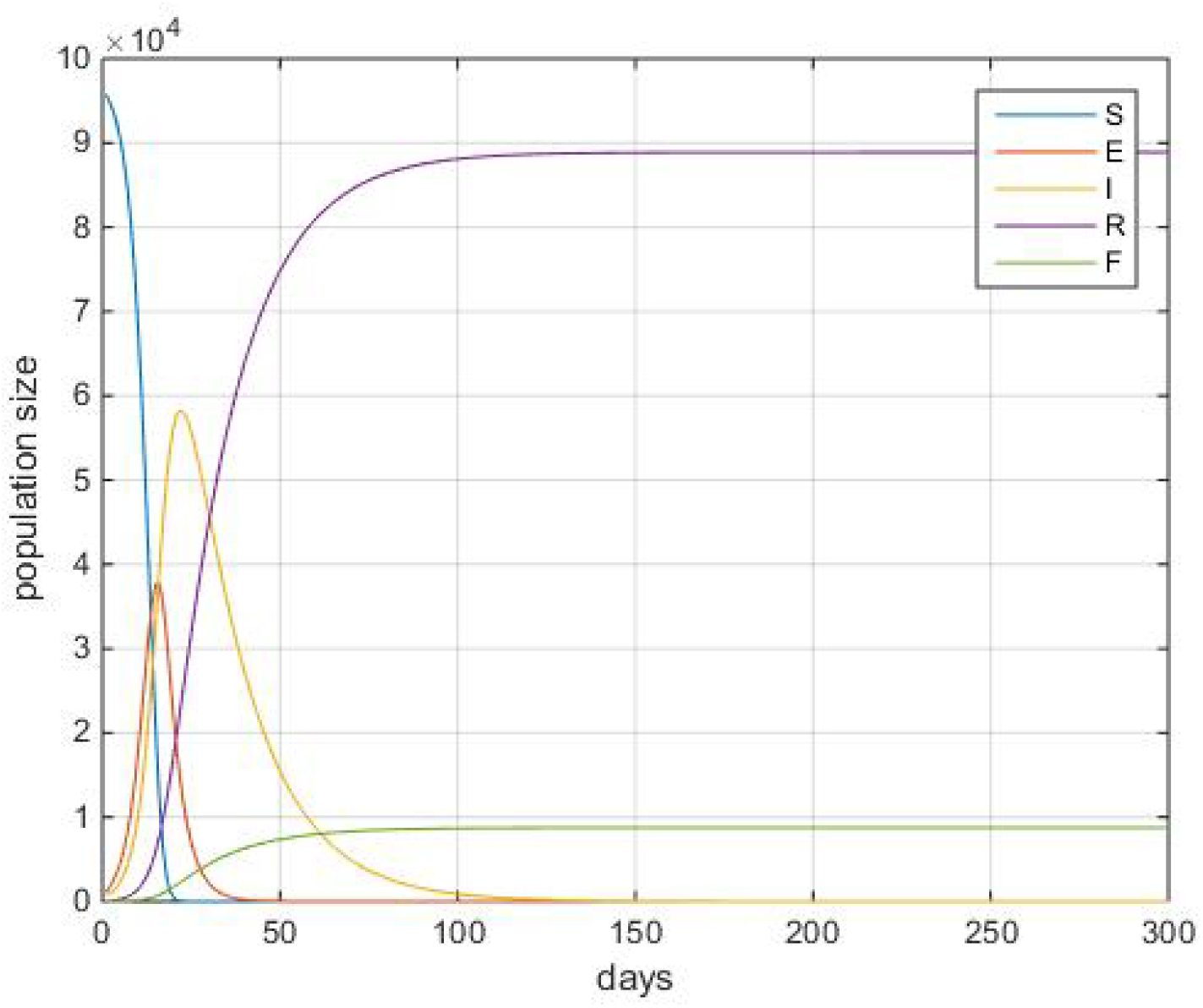
The main results of QSEIR model with the assigned values of parameters in baseline

Considering there have 20% estimation error of *errc*, the peak value of *I* would be in the range of [52,438, 64,090] and the peak date would be expected in the range February 7 to February 19, 2020. The end date would be in range from August 20, 2020 to September 1, 2020. During the period between March18-30, 2020 the epidemic would be totally controlled.

Yan *et al*. (2020) predicted that the peak value of confirmed cased in mainland China would be > 40,000, Hermanowicz (2020) predicted it to be 65,000, while Li and Feng (2020) estimated 51,600. There have been a number of studies estimating the peak number and date of confirmed cases in mainland China in the early stage of the epidemic (Batista, 2020; Gamero *et al*., 2020; Hermanowicz, 2020; Liu *et al*., 2020(a); Shi *et al*., 2020; Xiong and Yan, 2020). However, due to the limited emerging understanding of the new virus and its transmission mechanisms, their results were in the range from January 14, 2020 (Yan *et al*., 2020) to the beginning of March, 2020 (Geng *et al*., 2020) (see table 3). Most of them are in the mid of February, 2020, which are approximate to the real date February 17, 2020. The results of Wang *et al*. (2020b), Gamero *et al*. (2020) Xiong and Yan (2020), Li *et al*. (2020c), Hermanowicz (2020) and Shi *et al*. (2020) were in correspondence with our results, which are closer to the observed data. With 80% probability, our prediction of the peak date is 4 days ahead of the real date, the prediction error of the peak value is 0.43%, both estimates are much closer to the observed values compared with other published studies.

Furthermore, the existing studies seldom provided estimates of the duration of the epidemic and effects of different containment strategies in mainland China. At the regional level, Wu *et al*. (2020b) concluded that in Guangdong province, the epidemic would be totally controlled by mid to late March, 2020. The cumulative confirmed cases in Guangdong was ranked second among provinces in China. The number was 1,342 on February 22, 2020, which accounted for 1.74% of the cumulative confirmed cases in mainland China. The date on which no new exposed cases should be similar with that of mainland China. The result of Wu *et al*. (2020b) is correspondence with our result.

Yang *et al*. (2020) provided that in Chongqing the end date would be about May 11, 2020. The peak value of confirmed cases in Chongqing was 41 on January 30, 2020. The cumulative confirmed cases in Chongqing was 573 on February 22, 2020, accounting for 0.74% of the total in China. Therefore, its end date should be much earlier than that of mainland China. The end date of August 26, 2020 in mainland China in our research can be partly explained by Yang *et al*. (2020).

### Sensitivity analysis

How would *E, I, R, F* change if the value of parameters (*α*(*t*), *β*(*t*), *η*(*t*), *δ*(*t*), *γ*(*t*)) varied or if the beginning date January 23, 2020 of the simulation changed? We conducted sensitivity analyses of them in terms of their impacts on the *I* index one by one.

Figures 6-7 and tables 7-8 showed that the larger the value of *β*(*t*) *or δ*(*t*), the higher the peak value of the *I* index and the earlier the peak time. With the increase of *β*(*t*) *or δ*(*t*), their sensitive coefficient to *I* index decreased progressively. The sensitivity coefficient of *α*(*t*) to *I* index was the biggest. When *α*(*t*) increased 0.001%, 8,596 more confirmed cases will be observed (figure 10 and table 11). These results indicated that quarantine measures (or with vaccination that is not yet available) are the most effective containment strategy to control the epidemic. Figures 8-9 and tables 9-10 showed that the greater the value of *γ*(*t*) or *η*(*t*), the smaller the peak value of the *I* index. The peak date of *I* was not very sensitive to the change of *γ*(*t*). When *γ*(*t*) increased 1%, confirmed cases will be decrease between 4,395 and 7,432. When *η*(*t*) decreased 1%, 4,138 to 4,640 additional confirmed cases could be expected. The average absolute sensitive coefficient of *γ*(*t*) and *η*(*t*) to *I* ranked the second and third in those of five parameters (tables 7-11). This showed that to improve the rate of cure, the development of special medicine should be the second most effective measure.

**Table 7.** The peak value and peak date of *I* index when *β*(*t*) was changed and the sensitive coefficient of *β*(*t*) to *I*

| Value of $\beta(t)$ | Peak value | Peak date | The sensitive coefficient of $\beta(t)$ to $I$ |
| --- | --- | --- | --- |
| $\beta_1=1.2$ | 69915 | Feb.12 | / |
| $\beta_2=1.7$ | 72458 | Feb.9 | 5087 |
| $\beta_3=2.2$ | 73471 | Feb.7 | 2026 |
| $\beta_4=2.7$ | 74088 | Feb.6 | 1232 |
| $\beta_5=3.2$ | 74696 | Feb.6 | 1217 |

**Table 8.** The peak value and peak date of *I* index when *δ*(*t*) was changed and the sensitive coefficient of *δ*(*t*) to *I*

| Value of $\delta(t)$ | Peak value | Peak date | The sensitive coefficient of $\delta(t)$ to $I$ |
| --- | --- | --- | --- |
| $\delta_1=0.0474$ | 35986 | Mar.3 | / |
| $\delta_2=0.0948$ | 50463 | Feb.22 | 305423 |
| $\delta_3=0.1422$ | 59253 | Feb.17 | 185434 |
| $\delta_4=0.1896$ | 65353 | Feb.14 | 128712 |
| $\delta_5=0.2370$ | 69915 | Feb.12 | 96230 |

**Table 9.** The peak value and peak date of *I* index when *γ*(*t*) was changed and the sensitive coefficient of *γ*(*t*) to *I*

| Value of $\gamma(t)$ | Peak value | Peak date | The sensitive coefficient of $\gamma(t)$ to $I$ |
| --- | --- | --- | --- |
| $\gamma_1=0.0104$ | 93941 | Feb.15 | / |
| $\gamma_2=0.0208$ | 86197 | Feb.14 | -743216 |
| $\gamma_3=0.0313$ | 79944 | Feb.13 | -600016 |
| $\gamma_4=0.0417$ | 74495 | Feb.12 | -522950 |
| $\gamma_5=0.0521$ | 69915 | Feb.12 | -439580 |

**Table 10.** The peak value and date of *I* when *η*(*t*) was changed and the sensitive coefficient of *η*(*t*) to *I* index

| Value of $\eta(t)$ | Peak value | Peak date | The sensitive coefficient of $\eta(t)$ to $I$ |
| --- | --- | --- | --- |
| $\eta_1=0.0011$ | 71762.2 | Feb.12 | / |
| $\eta_2=0.0022$ | 71293.6 | Feb.12 | -426000 |
| $\eta_3=0.0032$ | 70829.6 | Feb.12 | -464000 |
| $\eta_4=0.0043$ | 70370.0 | Feb.12 | -417818 |
| $\eta_5=0.0054$ | 69914.8 | Feb.12 | -413818 |

**Table 11.**
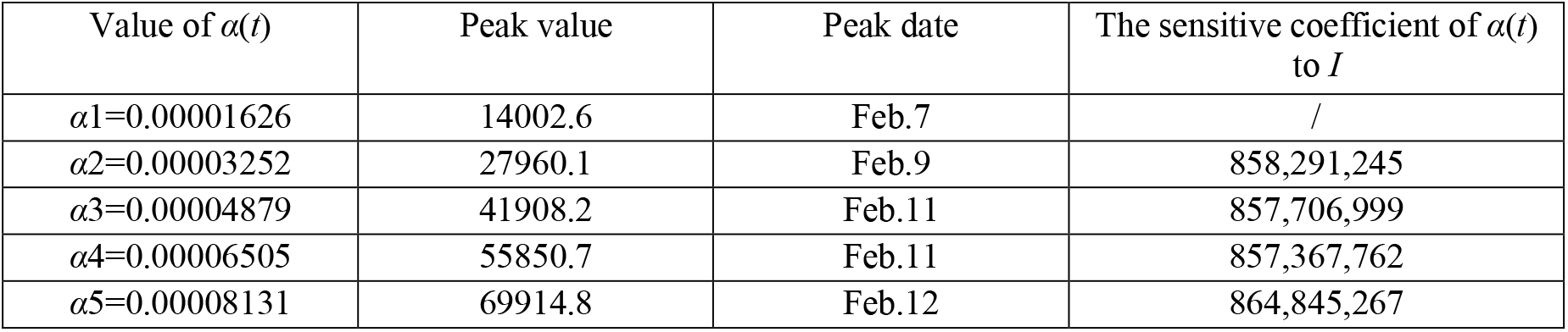
The peak value and date of *I* when *α*(*t*) was changed and the sensitive coefficient of *α*(*t*) to *I* index

**Figure 6.**
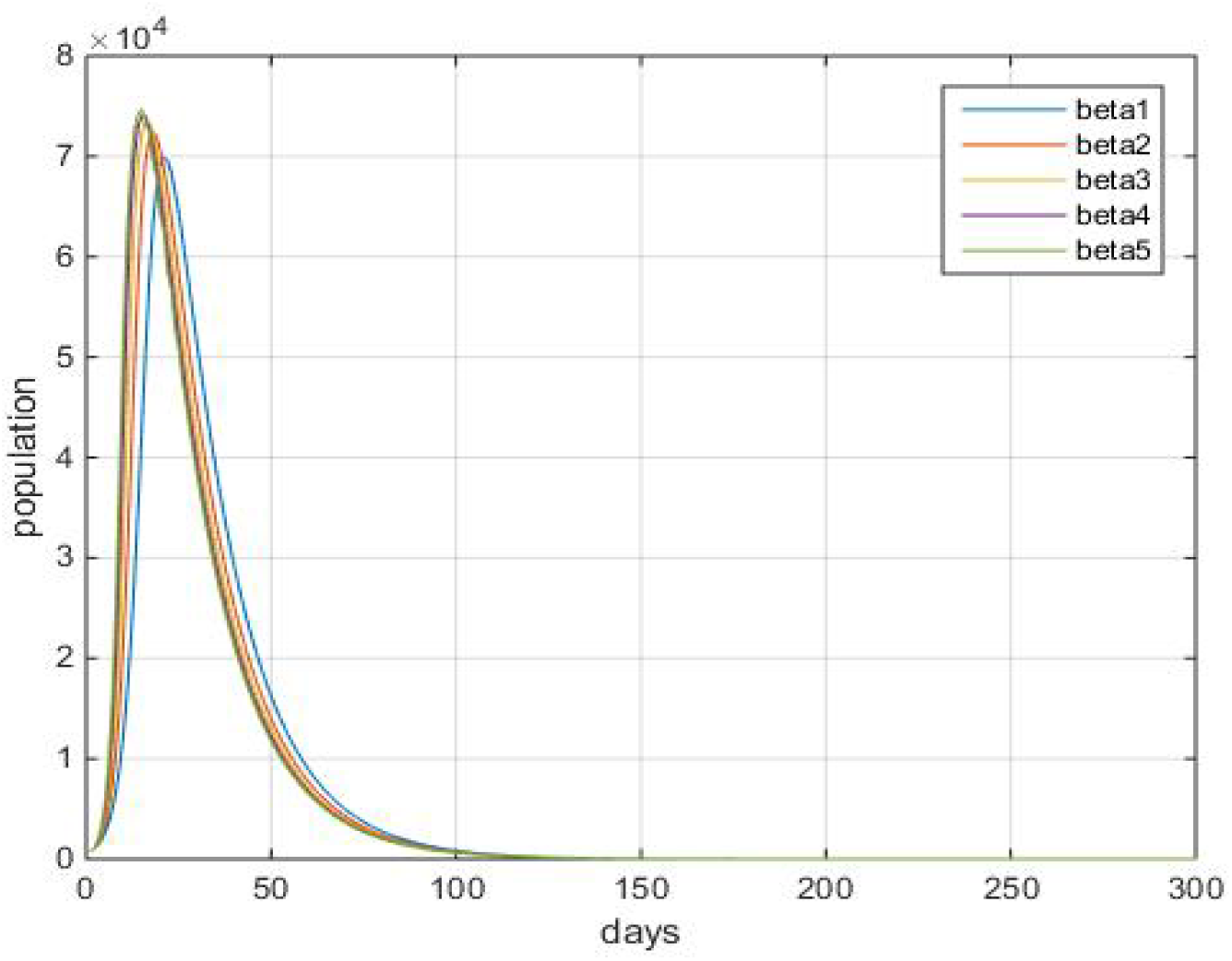
When *β(t)*=1.2 increase 0.5 each time and other parameters unchanged, the trend of *I* index

**Figure 7.**
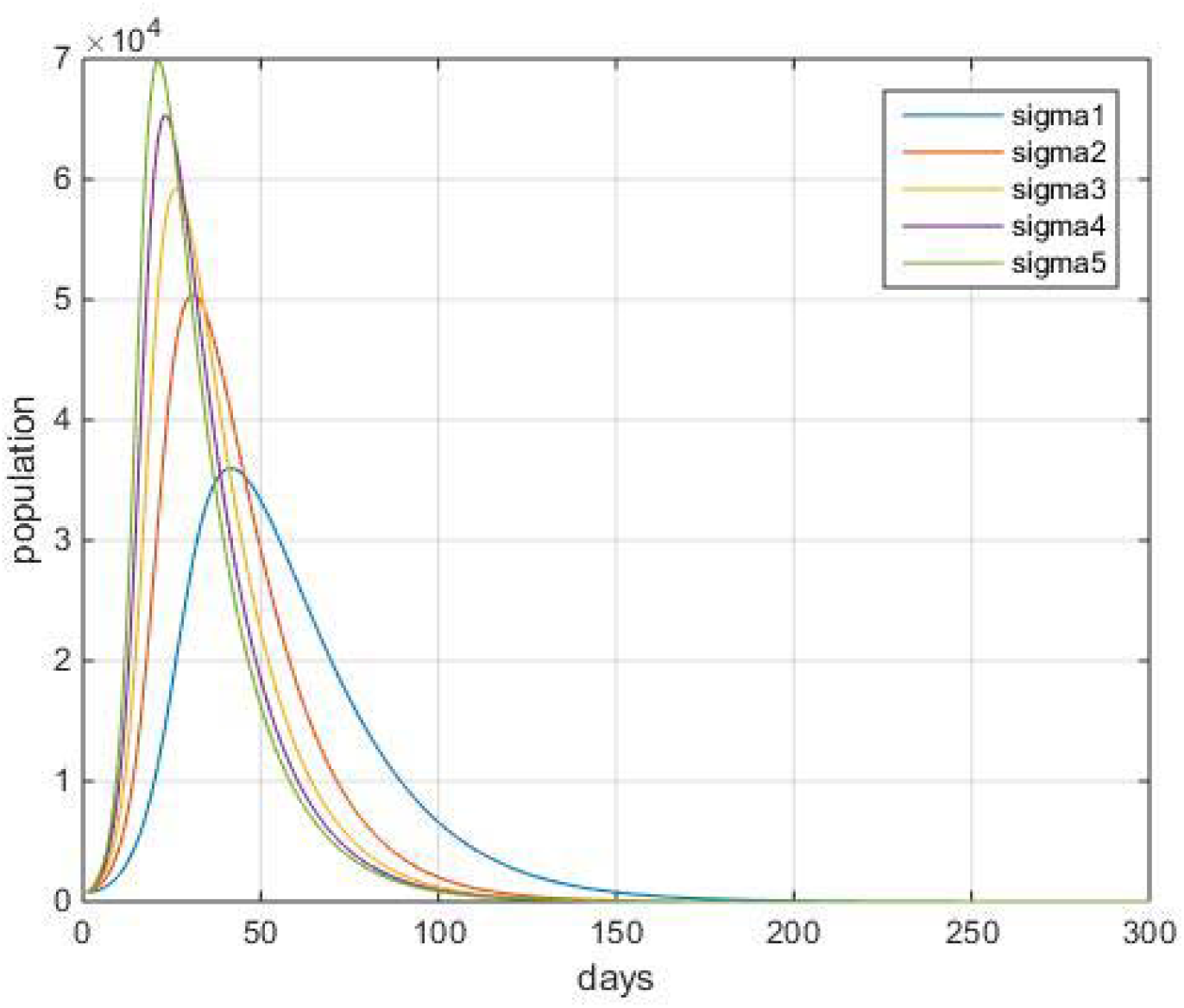
When *δ(t)*=0.2370 decrease 20% each time and other parameters unchanged, the trend of *I*

**Figure 8.**
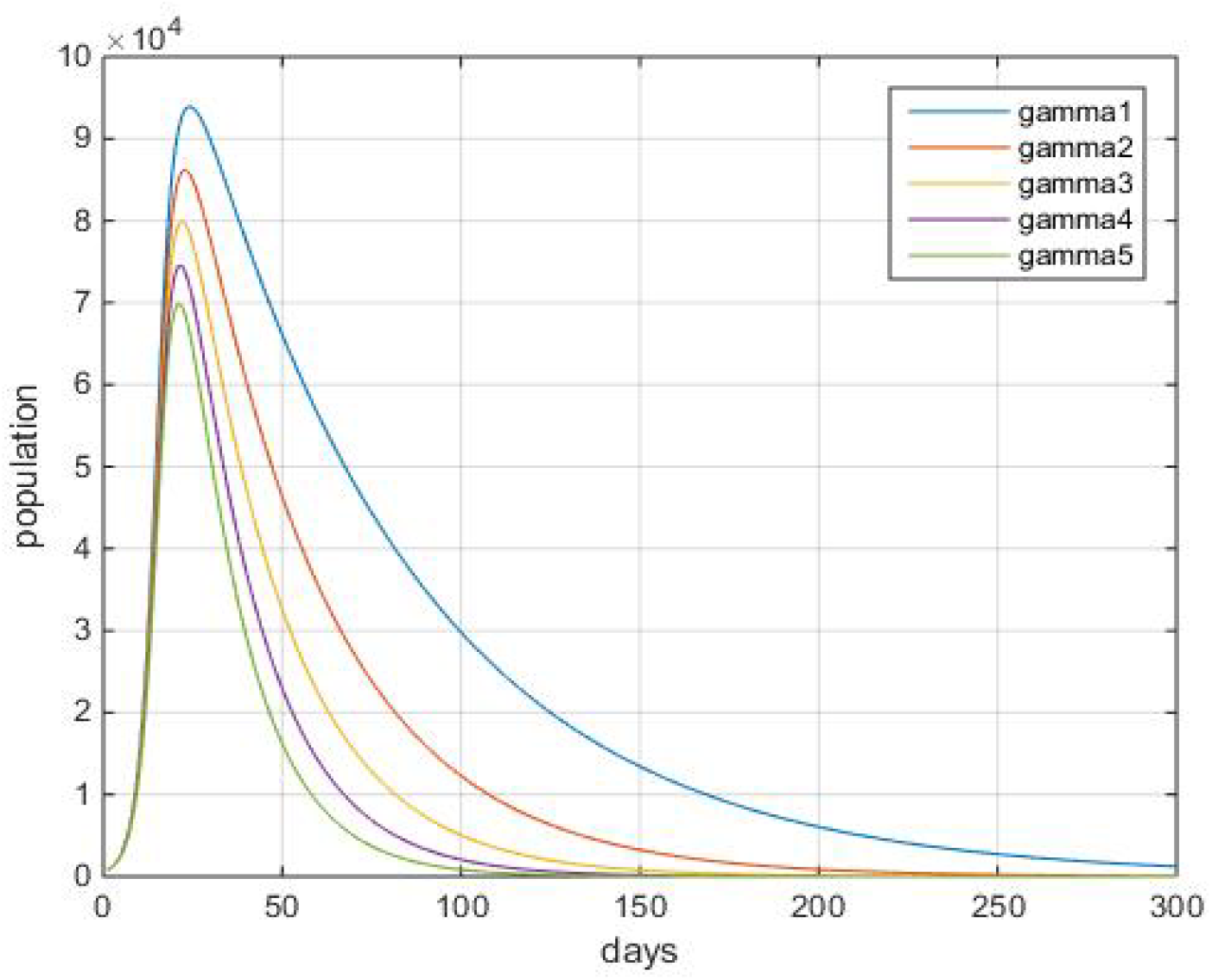
When *γ(t)*=0.0521 decrease 20% each time and other parameters unchanged, the trend of *I* index

**Figure 9.**
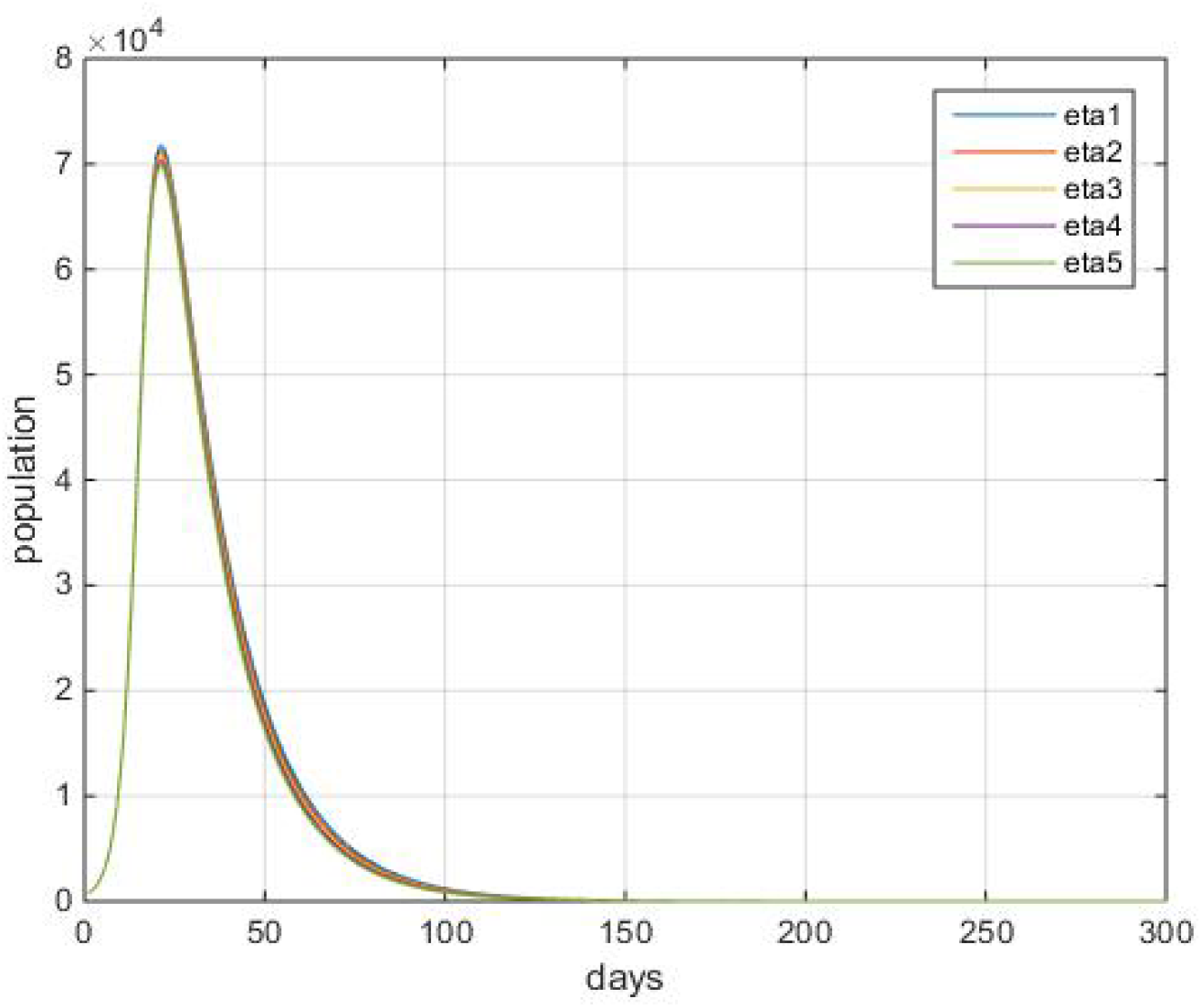
When *η(t)*=0.0054 decrease 20% each time and other parameters unchanged, the trend of I

**Figure 10.**
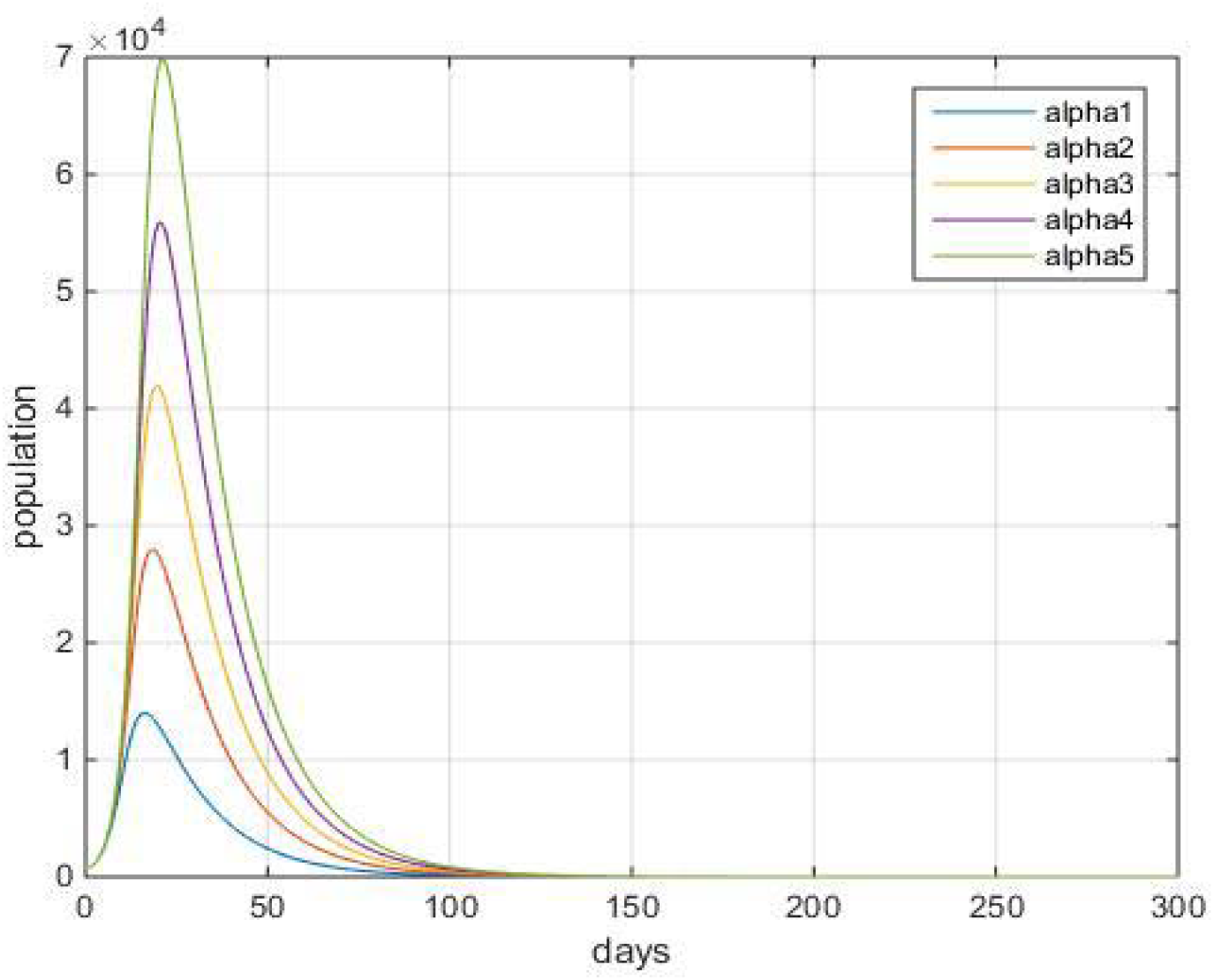
When *α(t)*=0.00008131 decrease 20% each time and other parameters unchanged, the trend of *I* index

If the beginning date of the simulation changed from January 23 to January 30 or February 6 in2020 with the value of variables in table 12, together with the same estimated value of parameters in table 5 and, QSEIR program, we can show the main results that started from January 30 in figure 11. Compared with the baseline, the peak value of the *I* index increased 0.9% or 1.5%. The peak date of *I* or the ended date of COVID-19 would be 3 days or 1 day ahead (figure 12 and table 12). Results mean that the simulating results were not sensitive to the initial start date. The QSEIR model system is stable.

**Table 12.** The initial values of variables when the simulation began from different dates and the main results

| Initial values |  |  |  |  | Main results |  |  |
| --- | --- | --- | --- | --- | --- | --- | --- |
| simulation began date in 2020 | $E$ | $I$ | $R$ | $F$ | peak value of $I$ | peak date of $I$ in 2020 | end date of the epidemic in 2020 |
| Jan.23 | 1072 | 771 | 34 | 25 | 69915 | Feb.12 | Aug.22 |
| Jan.30 | 15238 | 9308 | 171 | 213 | 70571 | Feb.11 | Aug.21 |
| Feb.6 | 26359 | 28985 | 1540 | 636 | 70953 | Feb.15 | Aug.25 |

**Figure 11.**
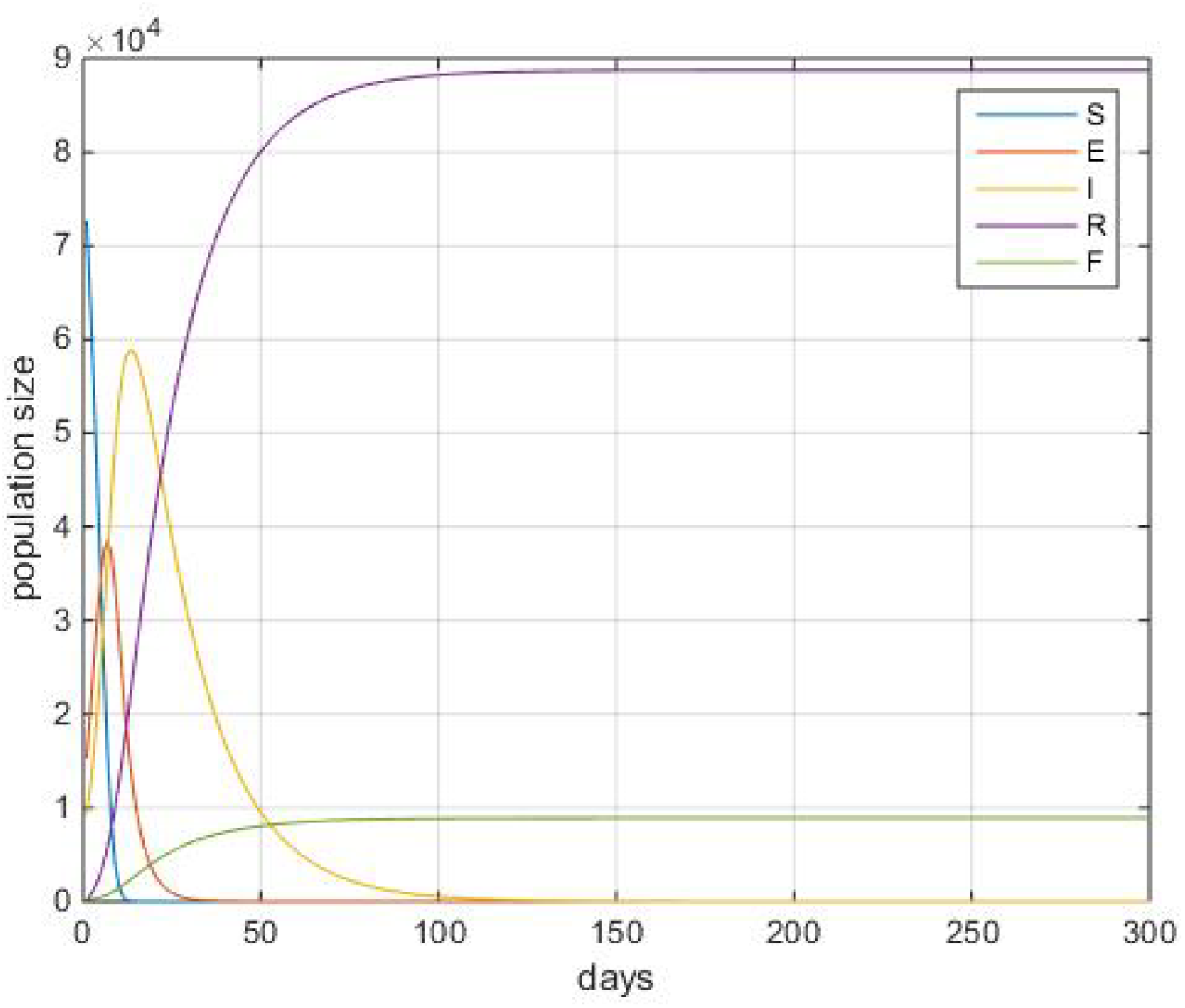
The main results of QSEIR model when the simulation began on January 30, 2020

**Figure 12.**
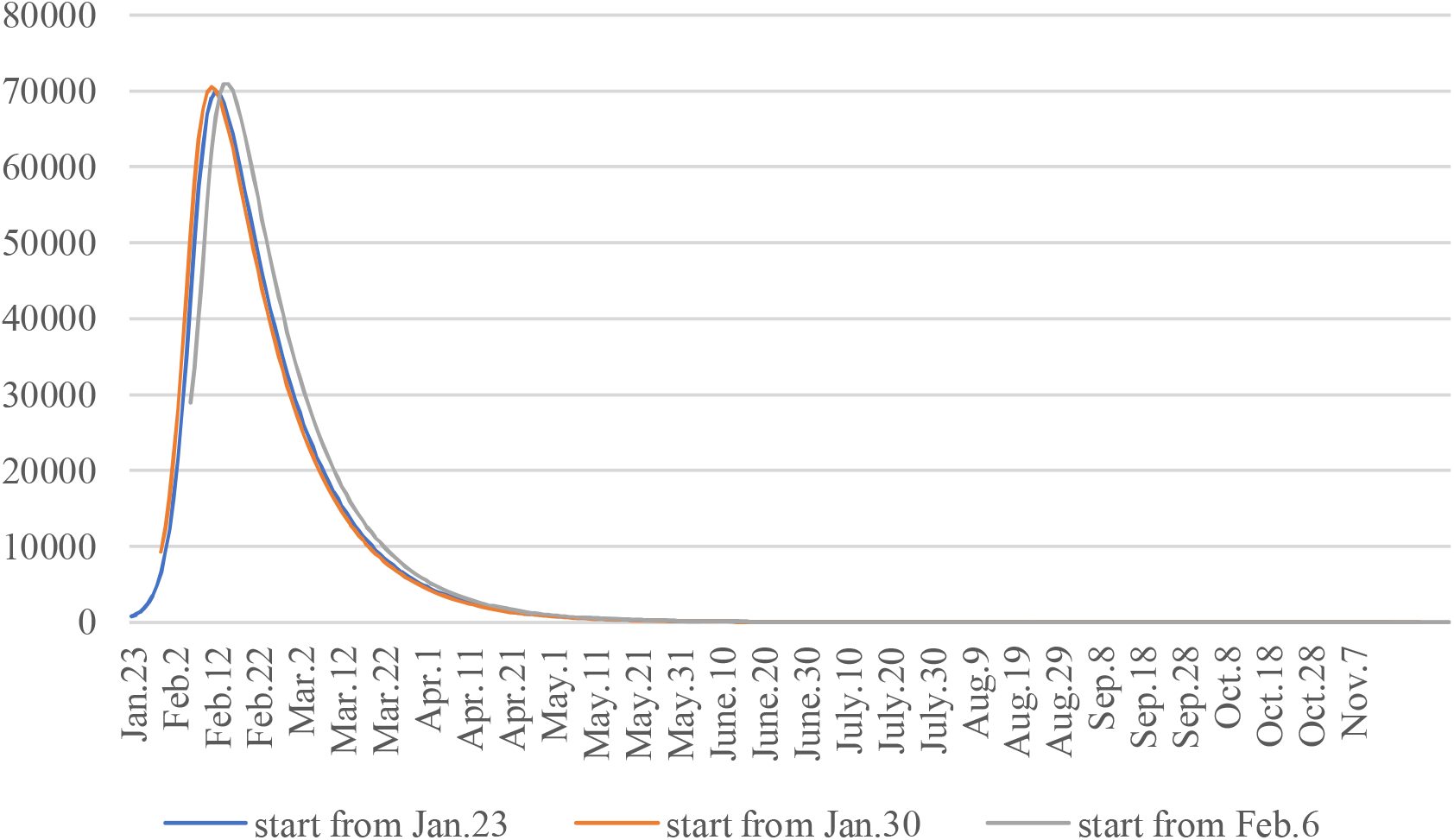
The comparison of *I* index when the simulation started from different dates

Due to the downward pressure on the economy, some enterprises resumed work one after another in compliance with the requirements of epidemic prevention and control. Because newly confirmed cases are decreasing day by day since February 17,2020, the outbreak was gradually brought under control, some people began to relax their vigilance. Some began to travel; some went out without masks. If the control measures are slightly relaxed from March 10, *α*(*t*) increased 0.00001 f rom 0.00006975, which means the number of *S* increased to 14,000, the end date would be exten ded from August 26, 2020 to September 14, 2020. And the date that the epidemic can be controlled would be extended 70 days, which would be on June 2^nd^, 2020. The cumulative confirmed cases would increase from 97,653 to 111,619, up 14.3% (figure 13). Evidence suggests that the colossal public health efforts of the Chinese Government have saved thousands of lives (Editorial, 20 20). It indicated that the quarantine measures should not be relaxed before the end of March, 202 0 in mainland China.

**Figure 13.**
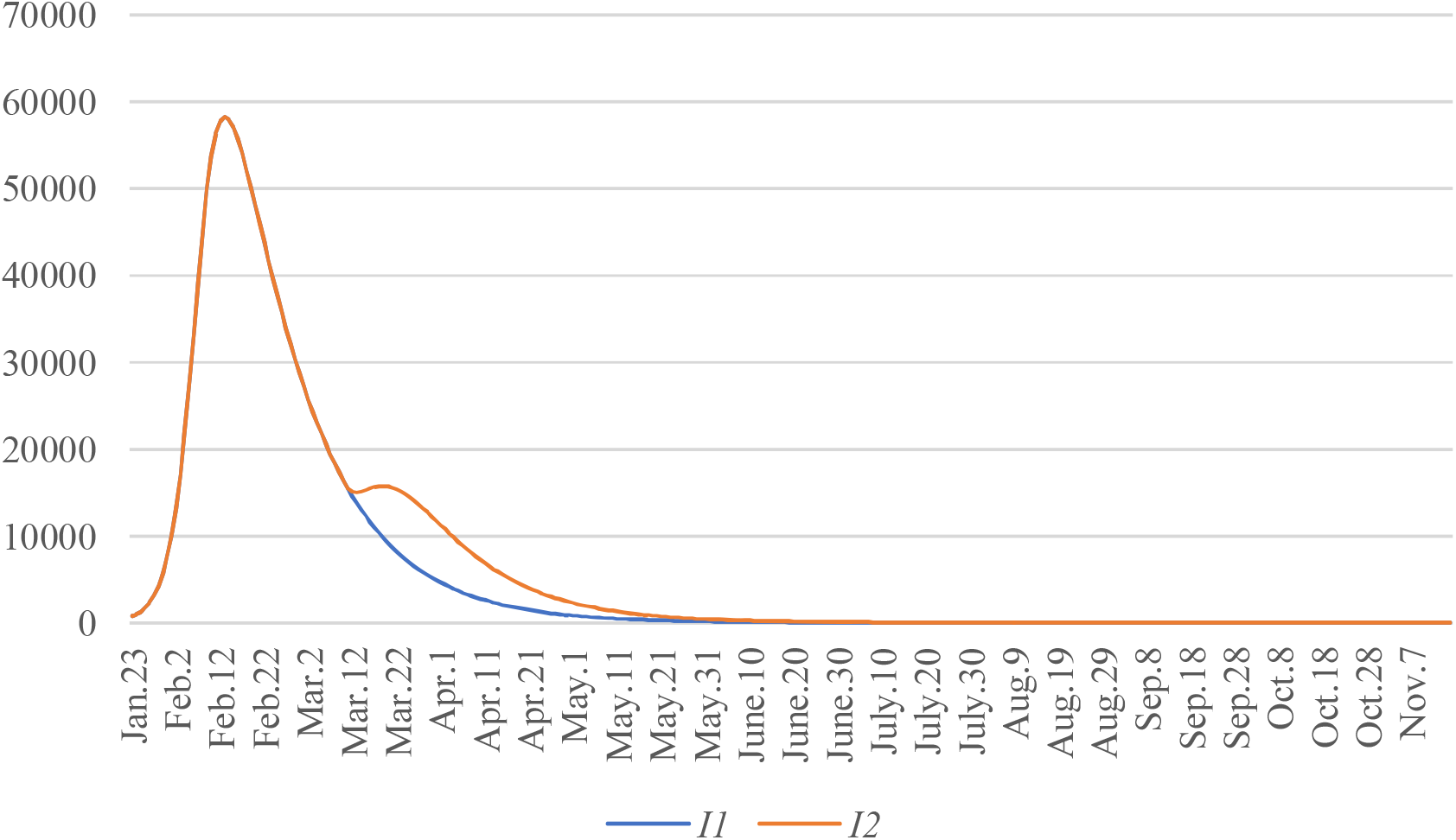
The comparison of *I* index when strict quarantine measures would be relaxed (alpha increased from 0.00006975 to 0.00007975) beginning on March10, 2020

## Conclusion

The paper proposed a QSEIR model that considers the unprecedented strict quarantine measures which are more fit for the epidemic situation in mainland China. Parameter estimation is the most critical part when using this kind of SEIR model to predict the trend of epidemic (Cao *et al*., 2020b). We estimated the model parameters reversely for the QSEIR model from published information with statistical methods and stochastic simulations; from these experiments, we found the parameters that achieved the best simulation test results. The application verified that the method is effective. The paper not only predicted the peak number and peak date of confirmed cases, but also provided estimates of the sensitivity of parameters of QSEIR, the duration of the epidemic and effects of different containment strategies at the same time. The long-term simulation result and sensitive analysis in mainland China showed that the QSEIR model is stable and can be empirically validated. It is suggested that the QSEIR model can be applied to predict the development trend of the epidemic in other regions or countries in the world.

## Discussions

In QSEIR model, the parameters are dynamically changing for each day. Parameters estimation is the most important part in the kind of SEIR model (Cao *et al*., 2020b). The paper illustrated the method to generate the parameter estimations. Given data limitation, we estimated a constant value to each of them with 20% errors in simulation tests, which was the best result in 50000 times stochastic simulation within their statistical ranges. We applied these values in prediction and obtained better results than existed researches. With the improvement of data quality and more data, variable parameters can be estimated and the forecasting accuracy of the model could be enhanced.

The vaccine research and development cycle are relatively long, from researching products to large-scale production and promotion, it takes about 6-18 months. It seems that the COVID-19 vaccination cannot be applied in large-scale quantities before the end of August, 20203.However, the COVID-19 is now spreading more seriously in other countries and regions in the world and there is also the possibility of its returning to China. As of March 7, 2020, 21, 110 confirmed cases of COVID-19 have been reported in 93 countries/territories/areas. Hence, it is imperative that the development of vaccines and specific drugs for COVID-19 should be promoted by many countries with the technical resources to conduct the necessary high-level research. Until they appear, it is the most important that appropriate quarantine measures are retained. In mainland China, the quarantine measures should not be relaxed before the end of March, 2020. China can fully resume production with appropriate anti-epidemic measures beginning in early April, 2020. The results of this study also implied that other countries now facing the epidemic outbreaks should act more decisively and take in time quarantine measures though it may have negative short-term public and economic consequences (Editorial, 2020).

## Data Availability

We collated epidemiological data from publicly available data sources (news, articles, press releases, and published reports from public health agencies). All the epidemiological information that we used is documented in the article.

http://www.nhc.gov.cn/xcs/yqtb/list_gzbd.shtml

## Contributors

Liu X L designed the QSEIR model, gave method to estimate parameters, compiled MATLAB program, got results and wrote the draft of the manuscript. Hewings G suggested to make sensitive analysis of parameters and estimate effects of different containment strategies. He edited the manuscript. Wang S Y explained some results and provided policy implications. Qin M H, Xiang X, Zheng S and Li X F collected data, some references and analyzed some data, the four of them made equal contributions to the paper.

## Declaration of interests

**We declare no competing interests**.

## Acknowledgements

This paper was supported by the 2019 Chinese Government Scholarship and National Natural Science Foundation of China under Grants No. 71874184 and No. 71988101.

## Footnotes

1 https://www.who.int/emergencies/diseases/novel-coronavirus-2019/situation-reports/

2 http://www.nhc.gov.cn/xcs/yqtb/list_gzbd.shtml

3 https://www.cnbeta.com/articles/science/947877.htm

## Notes

### Competing Interest Statement

The authors have declared no competing interest.

## References

1. Althaus C L. Estimating the reproduction number of Ebola virus (EBOV) during the 2014 outbreak in west Africa. PLoS Currents 2014; 6: ecurrents. outbreaks.91afb5e0f279e7f29e7056095255b288. doi:10.1371/currents.outbreaks.91afb5e0f279e7f29e7056095255b288.

2. Anastassopoulou C, Russo L, Tsakris A, et al. Data-based analysis, modelling and forecasting of the novel Coronavirus (2019-nCoV) outbreak. medRxiv 2020; doi: https://doi.org/10.1101/2020.02.11.20022186.

3. Ayittey F K, Ayittey M K, Chiwero N B, et al. Economic impacts of Wuhan 2019-nCoV on China and the world. Journal of Medical Virology 2020; 1–3. doi: https://doi.org/10.1002/jmv.25706.

4. Batista M. Estimation of the final size of the coronavirus epidemic by the logistic model. Feb 2020; doi: https://doi.org/10.1101/2020.02.16.20023606.

5. Bogoch II, Watts A, Thomas-Bachli A, et al. Pneumonia of unknown etiology in Wuhan, China: potential for international spread via commercial air travel. Journal of Travel Medicine 2020; doi:10.1093/jtm/taaa1008.

6. Cao Z, Zhang Q, Lu X, et al. Estimating the effective reproduction number of the 2019-nCoV in China. medRxiv 2020(a); doi: https://doi.org/10.1101/2020.01.27.20018952.

7. Cao Z, Zhang Q, Lu X, et al. Incorporating human movement data to improve epidemiological estimates for 2019-nCoV. medRxiv 2020(b); doi: https://doi.org/10.1101/2020.02.07.20021071.

8. CNN Business. Accessed February 9, 2020. https://edition.cnn.com/2020/01/31/economy/china-economy-coronavirus/index.html.

9. Cowling B J, Leung G M. Epidemiological research priorities for public health control of the ongoing global novel coronavirus (2019-nCoV) outbreak. Eurosurveillance 2020; doi: https://doi.org/10.2807/1560-7917.ES.2020.25.6.2000110.

10. Editorial. COVID-19: too little, too late? The Lancet 2020; 395, 755, March 7. doi: https://doi.org/10.1016/S0140-6736(20)30522-5.

11. Fan R G, Wang Y B, Luo M, et al. SEIR-based novel pneumonia transmission model and inflection point prediction analysis (in Chinese). Journal of University of Electronic Science and Technology of China 2020. http://kns.cnki.net/kcms/detail/51.1207.T.20200221.1041.002.html.

12. Gamero J, Tamayo J A, Martinez-Roman J A. Forecast of the evolution of the contagious disease caused by novel coronavirus (2019-nCoV) in China. arXiv preprint 2002.04739, 2020.

13. Geng H, Xu A D, Wang X Y, et al. Analysis of the role of current prevention and control measures in the epidemic of new coronavirus based on SEIR model (in Chinese). Journal of Jinan University (Natural Science & Medicine Edition) 2020; 41(2): 1–7. http://kns.cnki.net/kcms/detail/44.1282.n.20200214.1318.002.html.

14. Hartley D M, Smith D L. Uncertainty in SARS epidemiology. The Lancet 2003; 362:170–171.

15. Hermanowicz S W. Forecasting the Wuhan coronavirus (2019-nCoV) epidemics using a simple (simplistic) model. medRxiv 2020; doi: https://doi.org/10.1101/2020.02.04.20020461.

16. Huang Y P. Impact of new coronary pneumonia and policy responses (In Chinese). Business Observer 2020.

17. IHS Markit. Impacts of coronavirus containment effort ripple through global economy. Accessed February 9, 2020. http://news.ihsmarkit.com/prviewer/release_only/slug/.

18. IMF News. Statement by international monetary fund president Kristalina Georgyeva on the economic impact of the COVID-19 epidemic 2020 (In Chinese). https://www.imf.org/zh/News/Articles/2020/02/22/pr2061-remarks-by-kristalina-georgieva-to-g20-on-economic-impact-of-covid-19.

19. Joseph T W, Kathy L, Gabriel M L. Nowcasting and forecasting the potential domestic and international spread of the 2019-nCoV outbreak originating in Wuhan, China: a modelling study. The Lancet 2020. https://doi.org/10.1016/S0140-6736(20)30260-9.

20. Leung K, Wu J T, Leung G M. Nowcasting and forecasting the Wuhan 2019-nCoV outbreak. Preprint published by the School of Public Health of the University of Hong Kong 2020; https://files.sph.hku.hk/download/wuhan_exportation_preprint.pdf.

21. Li Q, Guan X, Wu P, et al. Early transmission dynamics in Wuhan, China, of Novel Coronavirus–infected pneumonia. New England Journal of Medicine 2020(a); doi: 10.1056/NEJMoa2001316.

22. Li W L, Zhang G L. Impact of new coronary epidemic on China’s economy (In Chinese). First Institute of Finance and Economics 2020. http://www.cbnri.org/news/5442298.html.

23. Li X, Wu Q, Benfu L. Can search query forecast successfully in China’s 2019-nCov pneumonia?. medRxiv 2020(b); doi: https://doi.org/10.1101/2020.02.12.20022400.

24. Lipsitch M, Cohen T, Cooper B, et al. Transmission dynamics and control of severe acute respiratory syndrome. Science 2003;300(5627): 1966–1970.

25. Liu Q, Liu Z, Li D, et al. Assessing the tendency of 2019-nCoV (COVID-19) outbreak in China. medRxiv 2020(a); doi: https://doi.org/10.1101/2020.02.09.20021444.

26. Liu T, Hu J, Kang M, et al. Transmission dynamics of 2019 novel coronavirus (2019-nCoV). bioRxiv 2020(b); doi: https://doi.org/10.1101/2020.01.25.919787.

27. Liu Y, Gayle A A, Wilder-Smith A, et al. The reproductive number of COVID-19 is higher compared to SARS coronavirus. Journal of Travel Medicine 2020(c); doi: 10.1093/jtm/taaa021.

28. Mills C, Robins J, Lipsitch M. Transmissibility of 1918 pandemic influenza. Nature 2004; 432: 904–906. https://doi.org/10.1038/nature03063.

29. Pastor-Satorras R, Castellano C, Mieghem PV, et al. Epidemic processes in complex networks. Review of Modern Physis 2015; 87(3): 925–979.

30. Rabajante J F. Insights from early mathematical models of 2019-nCoV acute respiratory disease (COVID-19) dynamics. arXiv preprint 2002.05296, 2020.

31. Razum O, Becher H, Kapaun A, et al. SARS, lay epidemiology, and fear. The Lancet 2003; 361: 1739–1740.

32. Read J M, Bridgen J R E, Cummings D A T, et al. Novel coronavirus 2019-nCoV: early estimation of epidemiological parameters and epidemic predictions. medRxiv 2020; doi: https://doi.org/10.1101/2020.01.23.20018549.

33. Richard H. Offline: 2019-nCoV outbreak—early lessons. The Lancet 2020; https://doi.org/10.1016/S0140-6736(20)30212-9.

34. Riou J, Althaus C L. Pattern of early human-to-human transmission of Wuhan 2019 novel coronavirus (2019-nCoV), December 2019 to January 2020. Eurosurveillance 2020, 25(4).

35. Sanche S, Lin Y T, Xu C, et al. The novel coronavirus, 2019-nCoV, is highly contagious and more infectious than initially estimated. arXiv preprint 2002.03268 2020.

36. Shen M, Peng Z, Xiao Y, et al. Modelling the epidemic trend of the 2019 novel coronavirus outbreak in China. bioRxiv 2020; doi: https://doi.org/10.1101/2020.01.23.916726.

37. Shi P, Cao S, Feng P. SEIR Transmission dynamics model of 2019 nCoV coronavirus with considering the weak infectious ability and changes in latency duration. medRxiv 2020; doi: https://doi.org/10.1101/2020.02.16.20023655.

38. Su S, Li X C, Hao H, et al. Advances in research on SARS-CoV-2[J/OL] (in Chinese). Journal of Xi’an Jiaotong University (Medical Sciences) 2020. http://kns.cnki.net/kcms/detail/61.1399.R.20200224.0944.010.html.

39. Tang B, Wang X, Li Q, et al. Estimation of the transmission risk of the 2019-nCoV and its implication for public health interventions. Journal of Clinical Medicine 2020; 9(2): 462. doi:10.3390/jcm9020462.

40. Tomie T. Understanding the present status and forecasting of COVID—19 in Wuhan. medRxiv 2020; doi: https://doi.org/10.1101/2020.02.13.20022251.

41. Tuite A R, Fisman D N. Reporting, epidemic growth, and reproduction numbers for the 2019 Novel Coronavirus (2019-nCoV) epidemic. Annals of Internal Medicine 2020.

42. Wallinga J, Teunis P. Different epidemic curves for severe acute respiratory syndrome reveal similar impacts of control measures. American Journal of Epidemiology 2004; 160: 509–516.

43. Wang H, Wang Z, Dong Y, et al. Phase-adjusted estimation of the number of Coronavirus Disease 2019 cases in Wuhan, China. Cell Discovery 2020(a); 6(1): 1–8. doi: https://doi.org/10.1038/s41421-020-0148-0.

44. Wang Z X, Liu Z and Liu Z J. 2019-nCoV analysis and forecast based on machine learning (in Chinese). Journal of Biomedical Engineering Research 2020(b). http://kns.cnki.net/kcms/detail/37.1413.R.20200213.0956.002.html.

45. WHO. Novel coronavirus (2019-nCoV) situation report - 7. Jan 27, 2020. https://www.who.int/docs/default-source/coronaviruse/situation-reports/20200127-sitrep-7-2019--ncov.pdf?sfvrsn=98ef79f5_2 (accessed Jan 27, 2020).

46. Guan W J, Ni Z Y, Hu Y, et al. Clinical characteristics of 2019 novel coronavirus infection in China. New England Journal of Medicine 2020, doi: 10.1056/NEJMoa2002032.

47. Wu J T, Leung K, Leung G M. Nowcasting and forecasting the potential domestic and international spread of the 2019-nCoV outbreak originating in Wuhan, China: a modelling study. The Lancet 2020(a); doi: https://doi.org/10.1016/S0140-6736(20)30260-9.

48. Wu W, Bai R H, Li D N, et al. Preliminary prediction of the epidemic trend of 2019 novel coronavirus (2019-nCoV) pneumonia in Guangdong province (in Chinese). Journal of Jinan University (Natural Science & Medicine Edition) 2020(b); 41(2): 1–6. http://kns.cnki.net/kcms/detail/44.1282.n.20200212.1132.004.html.

49. Xiong H, Yan H. Simulating the infected population and spread trend of 2019-nCov under different policy by EIR model. Available at SSRN 3537083 2020; doi: https://doi.org/10.1101/2020.02.10.20021519.

50. Yan Y, Chen Y, Liu K J, et al. Modeling and prediction for the trend of outbreak of NCP based on a time-delay dynamic system (in Chinese). Scientia Sinica Mathematica 2020; 50: 1–8. https://doi.org/10.1360/SSM-2020-0026.

51. Yang Y Q, Sun Q, Wang Y X, et al. Epidemic situation analysis and trend forecast of New Coronavirus Pneumonia (NCP) in Chongqing (in Chinese). Journal of Chongqing Normal University (Natural Science) 2020; 37(1). http://kns.cnki.net/kcms/detail/50.1165.n.20200218.0746.002.html.

52. Zhang B, Zhou H, Zhou F. Study on SARS-COV-2 transmission and the effects of control measures in China. medRxiv 2020; doi: https://doi.org/10.1101/2020.02.16.20023770.

53. Zhang J. Coronavirus will not cripple China’s economy, 2020. https://www.caixinglobal.com/2020-02-12/opinion-coronavirus-will-not-cripple-chinas-economy-101514545.html.

54. Zhao S, Lin Q, Ran J, et al. Preliminary estimation of the basic reproduction number of novel coronavirus (2019-nCoV) in China, from 2019 to 2020: A data-driven analysis in the early phase of the outbreak. International Journal of Infectious Diseases 2020; doi: https://doi.org/10.1016/j.ijid.2020.01.050.

55. Zhou T, Liu Q, Yang Z, et al. Preliminary prediction of the basic reproduction number of the novel coronavirus 2019-nCoV[J/OL] (in Chinese). Journal of Evidence-based Medicine 2020. http://kns.cnki.net/kcms/detail/51.1656.r.20200204.1640.002.html.

